# SARS-CoV-2 Orphan Gene ORF10 Contributes to More Severe COVID-19 Disease

**DOI:** 10.1101/2023.11.27.23298847

**Authors:** Jeffrey Haltom, Nidia S. Trovao, Joseph Guarnieri, Pan Vincent, Urminder Singh, Sergey Tsoy, Collin A. O’Leary, Yaron Bram, Gabrielle A. Widjaja, Zimu Cen, Robert Meller, Stephen B. Baylin, Walter N. Moss, Basil J. Nikolau, Francisco J. Enguita, Douglas C. Wallace, Afshin Beheshti, Robert Schwartz, Eve Syrkin Wurtele

## Abstract

The orphan gene of SARS-CoV-2, ORF10, is the least studied gene in the virus responsible for the COVID-19 pandemic. Recent experimentation indicated ORF10 expression moderates innate immunity in vitro. However, whether ORF10 affects COVID-19 in humans remained unknown. We determine that the ORF10 sequence is identical to the Wuhan-Hu-1 ancestral haplotype in 95% of genomes across five variants of concern (VOC). Four ORF10 variants are associated with less virulent clinical outcomes in the human host: three of these affect ORF10 protein structure, one affects ORF10 RNA structural dynamics. RNA-Seq data from 2070 samples from diverse human cells and tissues reveals ORF10 accumulation is conditionally discordant from that of other SARS-CoV-2 transcripts. Expression of ORF10 in A549 and HEK293 cells perturbs immune-related gene expression networks, alters expression of the majority of mitochondrially-encoded genes of oxidative respiration, and leads to large shifts in levels of 14 newly-identified transcripts. We conclude ORF10 contributes to more severe COVID-19 clinical outcomes in the human host.

## Introduction

As an orphan gene, the coding domain sequence (CDS) of ORF10 occurs only in severe acute respiratory syndrome coronavirus 2 (SARS-CoV-2), Pangolin-CoV-2019, and Bat-SL-CoV-RaTG13. The ORF10 CDS is absent from the genomes of other coronaviruses, viruses in general, and cellular organisms (1, 2).

Orphan genes (also called “ORFan” genes in viruses (3)) can arise from the de novo generation of new protein-coding sequences in nongenic regions of a genome, as novel ORFs within existing RNAs, or from the rapid large-scale modification of existing CDSs (4–7). If maintained during speciation, orphan genes can shape a phylogenetic lineage (8). This generation of new genetic elements provides a mechanism for the disruptive evolution of an existing trait or inception of a completely novel phenotype (5, 8–12).

Although the vast majority of viral, prokaryotic, and eukaryotic orphan genes are still unidentified (13), essential roles have been elucidated for many orphan genes (5, 12– 17). Orphan genes often affect phenotypes associated with ecological interactions, providing organisms with new opportunities for predation, parasitism, and defense (12, 17, 18), such as paralyzing toxins of parasitic wasps (19) and jellyfish (20). Orphan genes enable survival in freezing waters, and have evolved independently in numerous species (21).

Some orphan genes encode proteins that physically interact with transcription factors, altering gene expression and eliciting changes in traits that protect the host from biotic or abiotic stresses (22, 23), modify development (24, 25), or impact metabolism(23).

Initial reports suggested that ORF10 was neither transcribed or translated in the human host (26, 27). As such, in the scientific literature ORF10 was oddly belittled (ORF10 is “most peculiar, as it does not share sequence homology with any known protein” (28) and is “perhaps the least attractive of SARS-CoV-2 proteins” (29)), and is still often ignored, e.g. (30–32)

ORF10 does not appear to be necessary for SARS-CoV-2 transmission (28). However, abundant data shows that ORF10 can be both transcribed and translated (33–41). Furthermore, biochemical characterization of cell models expressing ORF10 indicate that ORF10 protein physically interacts with diverse human host proteins (1, 42–47). Targeted studies show that expression of ORF10 in cell models alters the cellular immune response in HEK293 cells via the mitochondrial antiviral signaling protein (MAVS) and degrades cilia in epithelial cells via the stimulation of interferon response cGAMP interactor 1 (STING1) (43, 47). These studies provide mechanistic insight on how ORF10 might act in cells. They highlight the importance of a direct evaluation of the effect of the ORF10 gene of SARS-CoV-2 in individuals with COVID-19.

To determine whether the ORF10 gene impacts the course of COVID-19 in the human host, we analyze SARS-CoV-2 genomes from five variants of concern (VOC) along with associated clinical disease data from GSAID (48, 49). Our results reveal that deviations from the canonical ORF10 sequence are associated with milder COVID-19 symptoms in humans. We globally evaluate ORF10 expression in the context of that of other SARS-CoV-2 genes and human host genes in RNA-Seq samples from diverse human tissues and disease stages. To gain broader understanding of the molecular events associated with ORF10, we identify genes and processes that are impacted by ORF10 expression in A549 and HEK293 human cell lines. Our results shed light on the 3D structure of the ORF10 protein and its 2D RNA structure, the evolutionary trajectory of ORF10, implicate ORF10 in COVID-19 severity, and reveal ORF10 transcription dynamics and its effects on transcription of host genes in human cell models.

## Results

### SARS-CoV-2 ORF10 sequences across genomes of five VOC

Early in the pandemic, ORF10 was described as the most highly conserved SARS-CoV-2 protein (50). To evaluate the evolution of the ORF10 sequence in the context of other SARS-CoV-2 genes, we determined the extent of mutations in SARS-CoV-2 genomes across the pandemic, through the emergence of the Omicron VOC. By investigating data from over three million SARS-CoV-2 genomes, made available at Fumagalli et al. (51), we gained insight into the SARS-CoV-2 hotspots for synonymous and non-synonymous mutations. ORF10 has the lowest level of non-synonymous mutations/genome/site, and one of the lowest levels of synonymous mutations/genome/site relative to the other SARS-CoV-2 genes (Fig. 1A). High rates of synonymous mutations/genome/site tend to occur in ORF3b, ORF6, and ORF7b, while ORF3b has an impressive 0.016 non-synonymous mutations/genome/site (Fig. 1A).

**Fig. 1.**
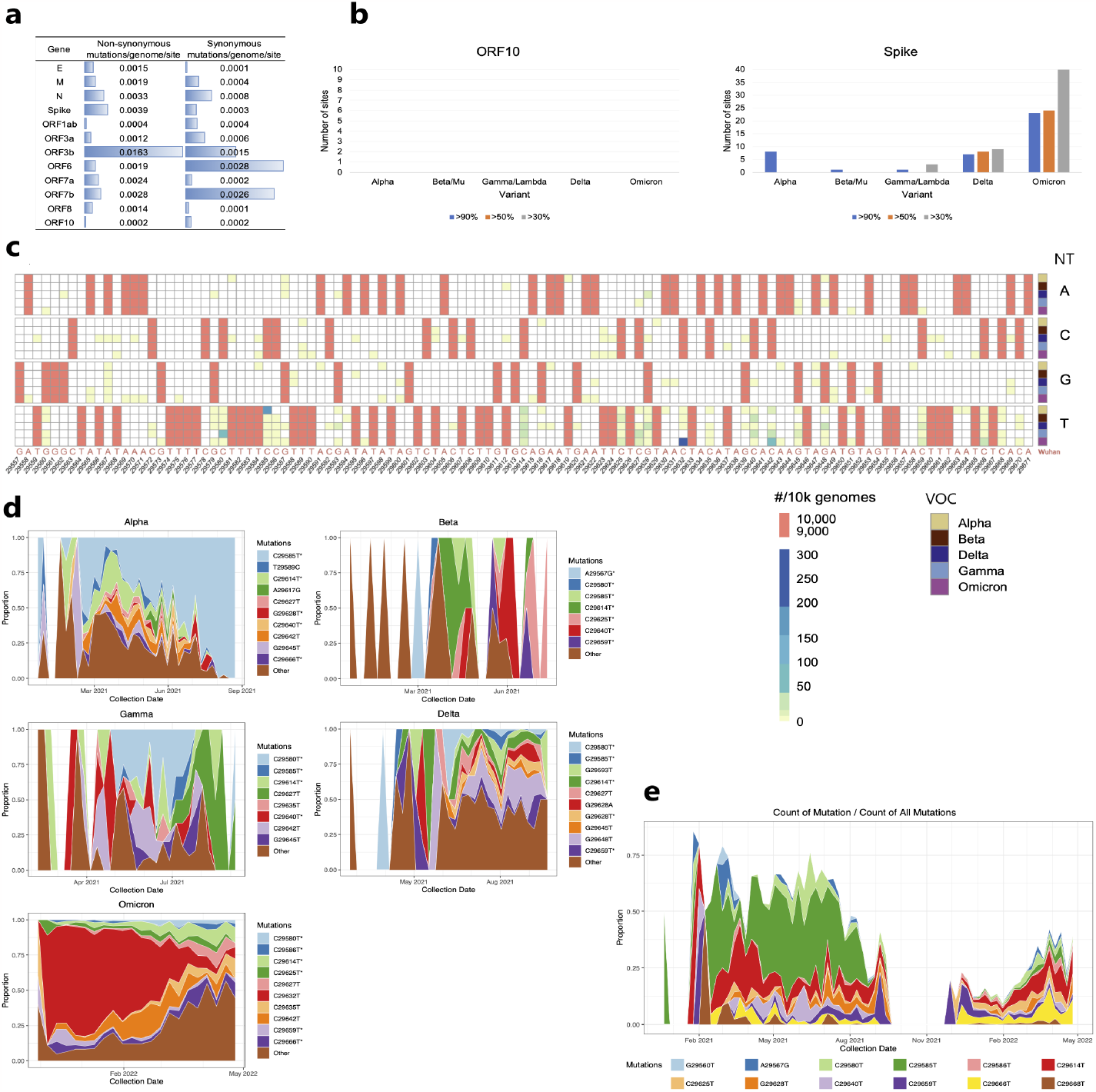
Paucity, skewness, and dynamics of ORF10 mutations in SARS-CoV-2 VOCs. All gene sequences are compared to those of the Wuhan-Hu-1 reference genome (GenBank NC_045512.2). **A**,**B**. Over three million SARS-CoV-2 genomes sampled in nasopharyngeal tissues during the pandemic (GSAID (48, 49, 51)). **C**,**D**,**E**. 210,101 ORF10 sequences with clinical metadata were retrieved from GSAID SARS-CoV-2 sequenced genomes (48, 49). **A**. ORF10 has the lowest number of nonsynonymous mutations/genome/nt site, and one of the lowest numbers of synonymous mutations/genome/nt site relative to other SARS-CoV-2 genes. **B**. Comparison of numbers of mutations in sequenced genomes for ORF10 and Spike protein of SARS-CoV-2. The ORF10 sequence has essentially remained identical across time and VOCs to the ORF10 Wuhan-Hu1 reference sequence. All sequences are compared to those of the Wuhan-Hu-1 reference genome. **C**. Mutations in ORF10 genetic sequence by VOC. Over 95% of ORF10 genes are identical in sequence to that of the Wuhan-Hu-1 strain. For comparability across VOC, the heatmap depicts data from 50^3^ randomly selected genomes (10^3^ from each VOC). Salmon colored font on the x-axis represents the original Wuhan-Hu-1 genetic sequence. NT, nucleotide in sampled sequences (if no substitution, nucleotide in X-axis will match nucleotide in NT-column); VOC, variant of concern. **D**. Mutations in VOC over time. Most ORF10 sequences were identical to ORF10 of the Wuhan-Hu-1 reference strain: Alpha VOC, 96% identical; Beta VOC, 99% identical; Delta VOC, 97% identical; Gamma VOC, 98% identical; Omicron VOC, 95% identical. The most prevalent mutations for each VOC are depicted. Y-axis, proportion of mutated sequences with a given mutation. The asterisks mark mutations that are present across all VOC. **E**. Co-circulation of ORF10 mutations over time. All VOC are included and the most prevalent mutations are depicted. Y-axis, proportion of mutated sequences with a given mutation. Terminology: “site” is used to indicate a specific location in a sequence; possible mutations at a given site are: the three possible substitutions (to a total of four nucleotides), an insertion, or a deletion; a “mutation” in a SARS-CoV-2 gene is defined as any sequence differing from the Wuhan-Hu-1 reference sequence.

We used these same data (51) to examine the extent of prevalent mutations in each SARS-CoV-2 gene across the pandemic. As anticipated, the greatest frequency of prevalent synonymous and non-synonymous mutations accumulate in the Spike gene, particularly during the Omicron VOC wave; specifically, >90% of genomes have 20 mutations in the Spike gene (Fig. 1B - right). This result is in stark contrast to the lack of any prevalent mutation in ORF10 (Fig. 1B - left). ORF1ab and N sequences significantly deviate from those of the Wuhan-Hu-1 strain, while all other SARS-CoV-2 genes had at least one prevalent mutation in one VOC, though ORF7a and ORF8 had reverted to the wild type Wuhan sequence by the Omicron VOC (Fig. 1A and Supplementary Fig. 1).

To investigate in more detail the ORF10 sequence over time and across VOC, and ultimately to determine whether ORF10 mutations are associated with COVID-19 severity, we assessed 210,101 SARS-CoV-2 genomes in GSAID for which there was associated clinical metadata (48, 49) (Supplementary Table 1).

Over 95% of ORF10 sequences were identical to that of the Wuhan-Hu-1 strain (Fig. 2D, Supplementary Table 1). The substitutions were distributed non-homogeneously throughout ORF10 (Fig. 1C). Fewer than 0.07% of ORF10 sequences had two or more mutations (Supplementary Table 1).

**Fig. 2.**
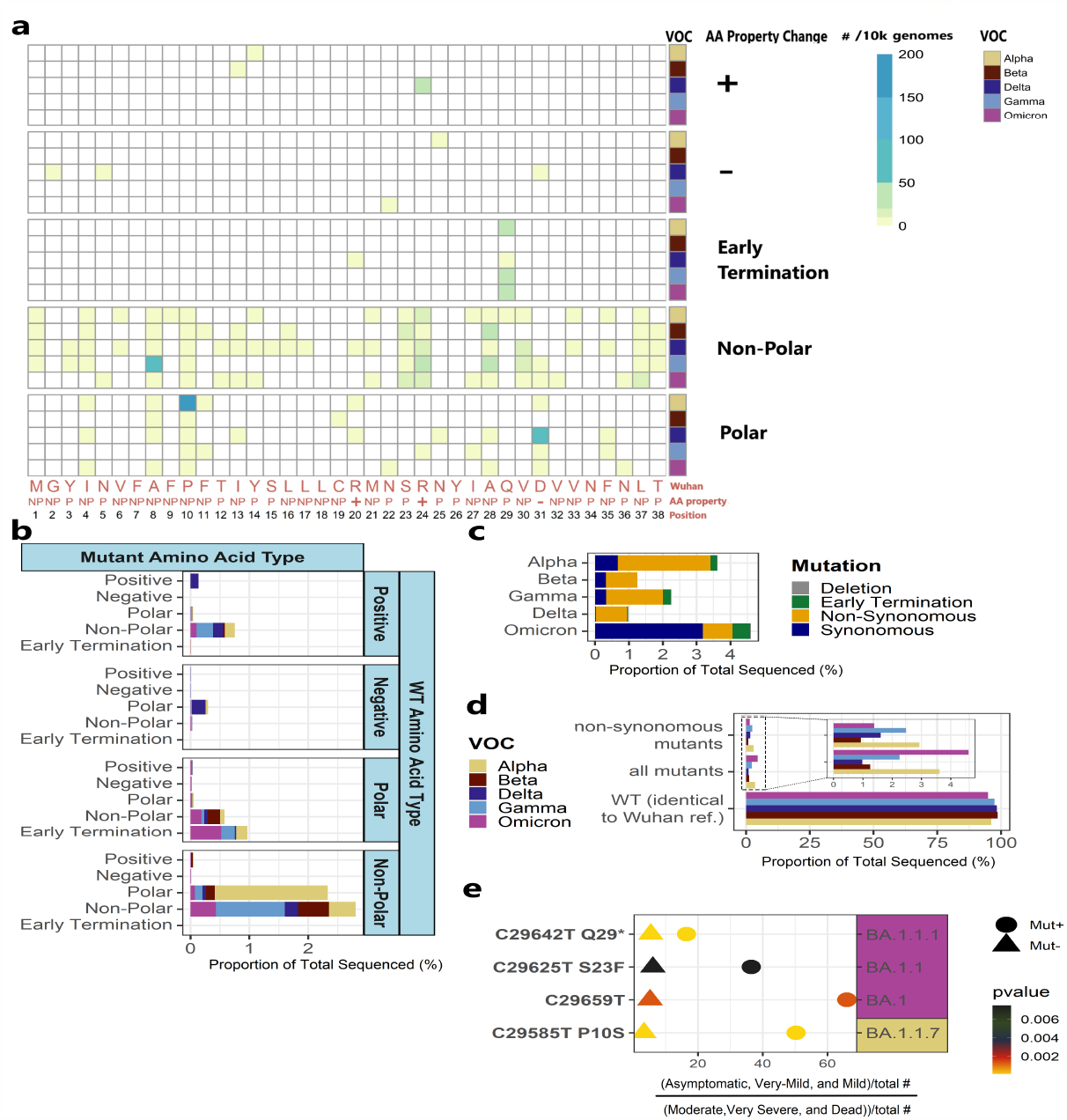
Mutations in SARS-CoV-2 ORF10 protein and their association with COVID-19 virulence. Data and metadata were retrieved from GISAID SARS-CoV-2 sequenced genomes (48, 49); 210,101 ORF10 sequences were aligned to the Wuhan-Hu-1 reference sequence. **A**. Non-synonymous AA changes in ORF10. Over half of the changes in the ORF10 protein are a substitution of one non-polar AA to a different nonpolar AA (in Beta, Gamma, Delta, and Omicron VOCs) or a polar AA (in Alpha VOC). For comparability across VOC, the heatmap depicts data from 50^3^ randomly sampled genomes, 10^3^ from each VOC. Salmon colored font on x-axis represents the original Wuhan-Hu-1 AA sequence and AA properties include (NP = nonpolar, P = polar, +/-depict charge). **B**. Non-synonymous AA changes in ORF10 by AA property. **C**. Proportions of synonymous, non-synonymous, deletion, and early termination mutations. Over two thirds of the changes in the Omicron VOC are synonymous substitutions, whereas in other VOCs most changes were non-synonymous substitutions. Deletions and early terminations are rare. **D**. Non-synonymous mutations relative to all mutations and Wuhan in each VOC. **E**. P-values for association of four ORF10 mutations with better clinical outcomes for patients with COVID-19. Outcomes were compared within each given VOC strain. Chi-square test was used to compare individuals with i) asymptomatic, very-mild, or mild symptoms, to individuals ii) with moderate, very severe symptoms or who died of this disease. These are: C29642T, which results in early stop codon Q29* in Omicron BA.1.1.1 (p-value = 2.84E-10); C29585T (P10S) in Alpha B.1.1.7 (p-value = 4.42E-32); and C29625T (S23F) in Omicron BA.1.1 (p-value = 7.64E-03). One synonymous mutation, C29659T in Omicron BA.1 (p-value = 1.29E-03). The x-axis normalizes the comparison.

Although C to T substitution bias is atypical of viruses in general, the phenomenon has been reported for genomes of Betacoronavirus species, including SARS-CoV-2 (52). C to T substitution is also a characteristic of humans (53). We examined the extent and distribution of C to T substitution bias in the ORF10 gene across the SARS-CoV-2 VOCs. C to T accounted for the majority of the substitutions (Fig. 1C and Supplementary Table 1). Surprisingly, the percentage of C to T substitutions in ORF10 differed substantially among the VOCs, ranging from 33% of all substitutions in Delta VOC to 89% of all substitutions in Omicron VOC. The majority of C to T substitutions were non-synonymous for each VOC except Omicron (Supplementary Table 1).

To compare among VOC, we randomly selected 10^3^ genomes from each VOC: Alpha, Delta, Omicron, Beta, and Gamma. In these 5x10^3^ genomes, some sites (i.e. 29601, 29605, 29610, 29634, 29656) are invariant from those of the wild type Wuhan-Hu-1 sequence. Most other sites are very rarely mutated (1-20 mutations per 10^3^ genomes) regardless of the VOC. Only four ORF10 mutations are present in more than 50*/*10^3^ genomes in any VOC (sites 29580, 29585, 29632, 29642) (Fig. 1C). Fewer than 0.03% of the sequenced genomes carry deletions within the ORF10 sequence; these deletions are clustered between positions 29574-29581; no insertional mutations were identified (Fig. 2C, Supplementary Fig. 2, Supplementary Table 1).

Each VOC has ORF10 mutations that dominated at different points during its respective epidemic (Fig. 1D,E). C29585T punctuated the Alpha VOC epidemic. Mutations C29625T and C29640T dominated the Beta VOC epidemics. Viruses belonging to the Gamma VOC had high frequencies of mutations at C29580T and C29627T. The Delta VOC wave was mainly marked by the G29648T mutation, whereas the C29632T mutation marked the first months of the Omicron VOC.

We sought to understand whether any of the prevalent ORF10 mutations were shared among the VOCs and, if so, to assess the extent of their simultaneous circulation (Fig. 1D,E). The locations of the mutations appear non-random. Notably, from January 2021 to August 2021, C29585T, the principal mutation seen in Alpha, also appeared in Beta, Delta, and Gamma. The two mutations that dominated the Alpha VOC wave (C29614T and C29585T) emerged in the first few months of the pandemic, and the C29614T mutation also dominated the first half of the Beta VOC wave. The second half of the Beta VOC is dominated by C29640T and, finally, C29625T. C29640T dominates the beginning of the Gamma VOC wave, and C29580T was the predominant mutation through the remainder of this wave. While Gamma VOC’s C29580T and Alpha VOC’s C29614T dominated the first months of the Delta VOC wave; subsequently, no mutations were highly dominant until the emergence of Omicron. The beginning of the Omicron VOC epidemic was dominated by C29659T, followed by a period where few shared mutations were circulating, and later (March and April 2022) showed an increase in the circulation of Omicron VOC’s C29666T and Alpha VOC’s C29614T (Fig. 1D,E).

A majority of non-synonymous changes in the ORF10 protein across most VOCs entailed a non-polar AA substituted to a different non-polar AA; however, in the Alpha VOC most changes were a substitution of a nonpolar AA to a polar AA (Fig. 2A,B). The ratio of synonymous/nonsynonymous mutations was small in all VOC except for Omicron, in which the number of synonymous mutations was over 2.7 times higher than the nonsynonymous (Fig. 2C). Several non-synonymous events were never or rarely detected among the ORF10 sequences. For example, in no genome were the two positively charged arginines mutated to encode a negatively charged amino acid (AA). In only three sequences was the negatively charged Asp altered to a positively charged AA (histidine) (2 in Delta, 1 in Omicron VOC) (Fig. 2A,B, Supplementary Table 1).

### Association of ORF10 mutations with clinical disease severity

To test the hypothesis that ORF10 contributes to COVID-19, we evaluated the association of the mutations with clinical severity in each major strain of the five VOCs, using a Chi-square analysis (Supplementary Table 2, Fig. 2E). Most non-synonymous and synonymous mutations occurred only a few times in a given strain, and thus did not have the statistical power to manifest any alterations in disease progression. However, several mutations with clinical metadata were present in sufficient numbers in a strain to enable us to potentially detect any association with clinical severity.

We grouped the clinical designation of individuals with COVID-19 into two groups: individuals that presented with asymptomatic or very mild to mild symptoms, and individuals with moderate to very severe symptoms or who died from the disease. This grouping resulted in 181,755 individuals with clinical data and sequenced SARS-CoV-2 genomes and strain designations whom we were able to study (Supplementary Table 3).

Four ORF10 mutations are significantly associated with a more positive disease outcome (p-value < 0.008) (Fig. 2E). Three of these four ORF10 mutations are non-synonymous: C29642T, which results in early stop codon Q29* in Omicron VOC BA.1.1.1 (p-value = 2.84E-10); C29585T (P10S) in Alpha VOC B.1.1.7 (p-value = 4.42E-32); and C29625T (S23F) in Omicron VOC BA.1.1 (p-value = 7.64E-03).

One synonymous mutation, C29659T in Omicron BA.1 (p-value = 1.29E-03), confers an improved outcome in COVID-19 progression.

Several other less prevalent mutations are associated with a more positive disease outcome (p-value < 0.05, full data in Supplementary Table 2). Interestingly, >98% of ORF10 sequences of Omicron VOC BA.1.16 bore the synonymous mutation C29632T. Thus, in this strain, there were insufficient ORF10 gene sequences identical to the wild type Wuhan-Hu1 to analyze effects on COVID-19 disease severity (Supplementary Table 3).

### Structural features of ORF10 protein variants associated with better clinical outcomes

The three-dimensional (3D) structure of the 38-residue ORF10 protein has not been experimentally determined. Therefore, we computationally predicted the 3D structure of the wild type ORF10 protein (Wuhan-Hu-1) and that of the three non-synonymous ORF10 mutant variants that are associated with improved patient outcomes (Fig. 2E), using RGN2 software (54). RGN2 implements a language and deep learning model that outperforms AlphaFold2 and RoseTTAFold for prediction of orphan gene protein structures.

The wild type ORF10 protein is predicted to fold into an α-helix consisting of alternating polar and nonpolar AA with charged side chains near the C-terminus (Fig. 3A). Mutation S23F swaps the polar AA, Ser, for the non-polar Phe. The amphipathic α-helix is maintained; however, the model indicates that the two Args are pushed apart, while the Asp flips direction (Fig. 3C). P10S causes Phe7 and all other AA before it to alter their direction (Fig. 3B). Electrostatic potential distribution shows a reduction of the area in the positive patch observed in the center of the helix for the S23F mutant, and an increase in the overall hydrophobicity of the α-helix when compared with wild type ORF10 (Fig. 3A and 3C).

**Fig. 3.**
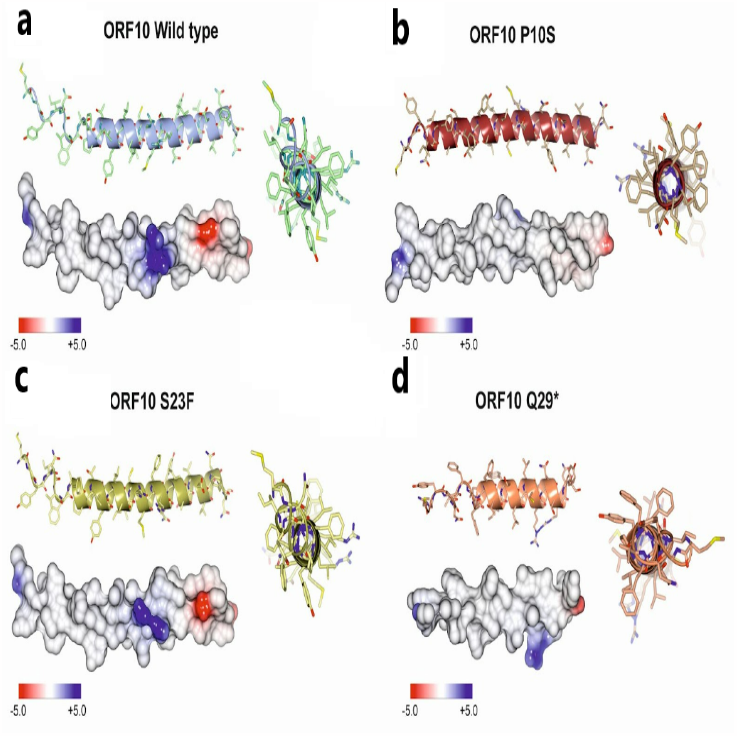
Structural features of the ORF10 protein and three mutants associated with reduced virulence. Structures of the protein encoded by the ORF10 wild type Wuhan-Hu-1 strain and those encoded by the three ORF10 non-synonymous mutants that were associated with a milder COVID-19 (see Fig. 2E) were modeled with RGN2 (54). Atomic coordinates were spatially aligned by the SSM superposition algorithm included in the Coot software (55). Each structure was represented using a ribbon model (side and top views) and the Van der Waals surface was colored by electrostatic potential (color scale below the surface model). **A**. ORF10 wild type Wuhan-Hu-1 reference genome; **B**. P10S ORF10; **C**. S23F ORF10; and **D**. Q29* ORF10.

Mutations that introduce a stop codon into ORF10, yielding a truncated protein, were extremely rare in any VOC. An exception is, the C29642T(Q29*) ORF10 variant, which occurs in 0.08-0.6% of Omicron, Alpha, Delta, and Gamma VOC sequences and is associated with a less severe clinical outcome. C29642T(Q29*) results in premature termination of the ORF10 protein just past the second Arg residue, leading to a shortened amphipathic α-helix missing the negatively charged Asp, leaving only positive amino acids. A second premature termination mutant, ORF10 R20*, was present solely in 2 sequences of the Omicron VOC (Supplementary Table 2); patients with this variant allele had mild symptoms, but sample numbers are insufficient to determine statistical significance. Our finding extends the data of (28); these researchers identified two patients, both with cases of mild COVID-19, who were infected with a truncated SARS-CoV-2 ORF10; they concluded that ORF10 is not required for SARS-CoV-2 replication (28).

### Structural features of wild type ORF10 RNA and its clinically-relevant mutant allele

We computationally predicted the structure and dynamics of wild type ORF10 RNA. We used ScanFold, a software developed to model significantly stable RNA secondary structures and used to develop a database for structures of genes of several viruses, including SARS-CoV-2 (56). We queried the ScanFold database predictions at https://structurome.bb.iastate.edu/sars-cov-2 to obtain structural predictions on SARS-CoV-2 ORF10 (Fig. 4).

**Fig. 4.**
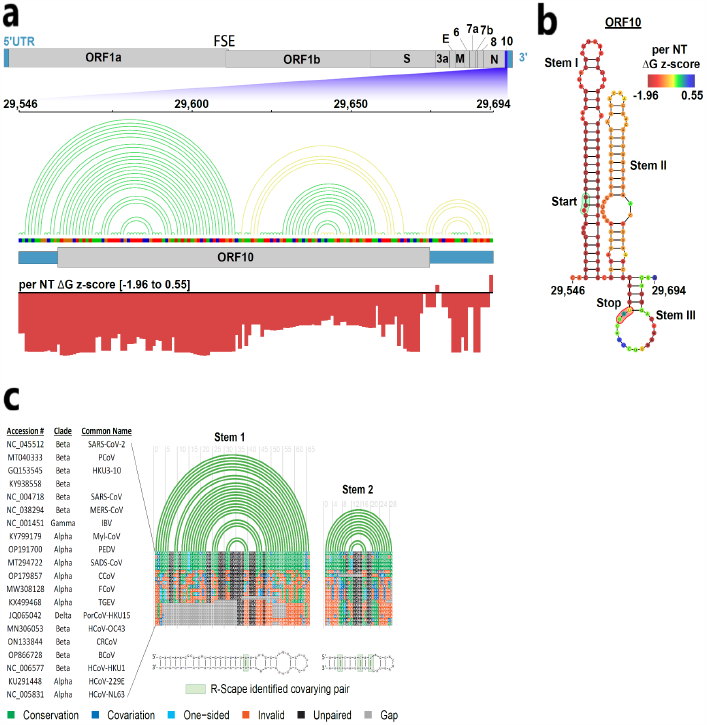
Secondary structure analysis of SARS-CoV-2 ORF10 RNA and its conservation in Betacoronaviruses. ORF10 wild type Wuhan-Hu-1 sequence is shown. **A**. Top, a representation of the SARS-CoV-2 genome with the 5- and 3-untranslated regions colored in teal and the coding regions colored in gray. ORF10 is highlighted in purple and a zoomed-in representation of the region from nucleotides 29546 to 29694 is shown. The 2D base pairing of this sequence is represented by the colored arc diagram where green arcs represent BPs with ΔG z-scores lower than - 1 and greater than - 2 and yellow arcs represent ΔG z-scores greater than - 1. Below, arc diagram of a sequence track of nucleotides; A (green), C (blue), U (red), and G (orange). Below there is a representation of the ORF10 coding region (gray) flanked by the untranslated sequence (teal). Red bars represent a per nucleotide (NT) ΔG z-score track of the region. **B**. 2D representation of the ORF10 RNA structure. The per NT ΔG z-score is overlaid on each nucleotide and the heatmap scale is annotated and shown (top right). The ORF10 start codon is labeled and annotated in green, the stop codon in red. Each major stem is labeled Stem I, II, or III based on the order they appear. **C**. RNA structure is conserved between the SARS-CoV-2 ORF10 and other species of Betacoronavirus. R-Chie arc diagrams for the two ScanFold predicted hairpins are shown with conservation annotated on an alignment of human coronaviruses. Accession numbers, genera, and common names for each aligned sequence are indicated on the left. All base pairs of ORF10 are 100% conserved among the four most closely related Betacoronavirus genomes (green). Consistent (single point) mutations (light blue) and compensatory (double point) mutations (dark blue) that preserve structure are found in related Betacoronaviruses. Nucleotides that do not maintain SARS-CoV-2 ORF10 structure are denoted in orange and gaps in sequence alignments are denoted in gray. Below the R-Chie arc diagrams are 2D representations of each hairpin, with consistent and compensatory mutations indicated (2D representations visualized with VARNA). Statistically significant covariation sites identified by R-Scape are annotated with green boxes on the 2D models.

Overall, the ORF10 region is inferred to be structured, with most nucleotides participating in base-pairing that provides ordered stability. Three stem-loops are predicted for the region encompassing ORF10, with the first two hairpins being larger and having more significant stability (i.e. a low ΔG z-score). The average per nucleotide ΔG z-score of this region is - 1.19, with a minimum and maximum value of - 1.96 and 0.55, respectively. Negative z-scores indicate the number of standard deviations more stable than random in a native RNA sequence, hence, ORF10 has more non-random sequence order and, therefore, a potential for function affected by RNA secondary structure.

The lowest z-score nucleotides occur in the first large stem, while the second stem contains more moderate yet predominantly negative z-score nucleotides. Both of the large stems have stretches of continuous base pairs punctuated with bulges, internal loops, and terminal hairpins, with the start codon for ORF10 occurring partway in the bulge on the 5’-end of the first stem (Fig. 4A,B).

The final, small stem, which encompasses the stop codon, has the most positive (i.e., least significant) z-score of the ORF10 gene. The collection of higher z-scores in this stem indicates that the RNA structure may be less likely to play a functional role than the upstream stem loops, and may be transient or dynamic (Fig. 4A,B).

To determine whether the ORF10 RNA itself might have consistent structural features, we evaluated the conservation of the structure of the ORF10 region in diverse coronaviruses. R-Chie arc diagrams for the two ScanFold-predicted hairpins are shown with conservation annotated on an alignment of diverse coronaviruses (Fig. 4C). Both hairpins are 100% conserved in the *structure* between human (SARS-CoV and SARS-CoV-2), bat (GQ153545 and KY938558), and pangolin (MT040333) strains of the SARS coronaviruses. Coronavirus genomes of Alpha-, Delta-, and Gamma coronavirus genera are more distantly related to SARS-CoV-2 (a Beta-coronavirus); these share little sequence or structural similarity to ORF10.

A query of SARS-CoV-2 ORF10 against all Coronaviridae sequences in the ViPR database (https://www.viprbrc.org/brc/home.spg?decorator=vipr) for evidence of covariation, to assess concerted evolution of paired sites that would preserve RNA structure, indicates the first two hairpins have statistically significant covarying base pairs (Fig. 4C). Almost all of the base pairs in the top five alignment tracks are 100% conserved, being either identical, consistent (single point) mutations, or compensatory (double point) mutations that preserve structure. For example, in the second, shorter hairpin, a CG base pair occurs as a compensatory AU pair in bat and SARS-CoV sequences, and as a UA pair in pangolin; a UA pair occurs as a compensatory CG base pair in most other closely conserved sequences. In general, structure is conserved in human, bat, and pangolin Betacoronavirus strains, with more distant sequences/strains unable to form stable structures with similar base pairing. This analysis is supportive of the model structure; the high conservation of the ORF10 RNA structure in Betacoronaviruses that lack an open reading frame to encode the ORF10 protein (Fig. 4) is indicative that ORF10 RNA itself might have a biological function in these viruses.

The synonymous ORF10 mutation, C29632T, predominated the first four months of the Omicron VOC wave (Fig.1D) and was associated with a better clinical outcome (Supplemental Table 2). We evaluated the effect of this mutation on the secondary structure of ORF10 RNA using ScanFold. C29632T converted a CG base pair to a UG wobble base pair. The mutation did not impact the modeled secondary structure according to ScanFold. Interestingly, the local thermodynamics of the ORF10 transcript were significantly affected. The mutation caused changes in the predicted minimum free energy (MFE) structure and positional entropy of the nucleotides around the mutations. Positional entropy, calculated by the RNAfold program, is a measure of a nucleotide’s likelihood of being in a specific conformational state. A low entropy indicates greater certainty in the model structural arrangement of a nucleotide, while a high entropy indicates less certainty, as alternative arrangements have potential for formation. The MFE values for the wild type and synonymous mutation sequences were similar, with the C29632T mutation having MFE (−31.6 kcal/mol). The average positional entropy for the wild type and C29632T sequences were 0.59 and 0.78, respectively. The C29632T mutation increased the average positional entropy of the sequence compared to the wild type.

The conservation of secondary structure is consistent with a functionality for ORF10 RNA. A functionality of ORF10 RNA would entail expression of the transcript. To our knowledge, expression of ORF10 RNA in any Betacoronavirus other than SARS-CoV-2 had not been reported. We analyzed ORF10 transcript levels in samples of intestinal organoids that were exposed to SARS-CoV Betacoronavirus, which does not contain an ORF10 open reading frame (57). ORF10 RNA is highly accumulated in these SARS-CoV-infected samples, providing further evidence consistent with a role for ORF10 RNA in Betacoronavirus pathology.

### SARS-CoV-2 ORF10 transcript is dis-coordinately accumulated across tissues of COVID-19 patients and SARS-CoV-2-infected organoids and cells

To determine the extent of ORF10 expression, and to gauge whether developmental, genetic, or environmental conditions might influence the accumulation of SARS-CoV-2 ORF10 transcripts relative to other SARS-CoV-2 transcripts, we analyzed levels of SARS-CoV-2 transcripts in raw data from RNA-Seq data representing 2,070 diverse SARS-CoV-2-associated human tissues and cells from 60 independent studies downloaded from Sequence Read Archives (SRA, https://www.ncbi.nlm.nih.gov/sra). We anticipated that this analysis might also shed light on reports (e.g., (26, 27)) that ORF10 is not transcribed. Samples include nasal swabs, blood cells and autopsied organs from individuals with COVID-19, and, *in vitro* SARS-CoV-2-infected human organoid models, primary, and established cell lines.

This analysis of the raw data provided a repertoire of information on SARS-CoV-2 transcript accumulation, most not quantified previously (Fig. 5A, Supplementary Files 1,2; Supplementary Fig. 3). SARS-CoV-2 transcripts, including ORF10, were abundant in most of the hundreds of samples from the viral portal of entry in human nasal tissues, and particularly abundant in individuals with high-viral titres; in contrast, SARS-CoV-2 transcripts were undetectable in the 1079 samples of human blood of individuals with COVID-19 (Fig. 5A, Supplementary Fig. 3). However, few samples from heart, liver, brain, kidney, bowel, or lymph nodes of COVID-19 autopsy patients had detectable SARS-CoV-2 transcripts (3/42 heart samples, 4/37 liver samples, 0/9 frontal cortex samples, 1/18 mediastinal lymph node samples, 0/30 kidney samples, and 1/4 bowel autopsy samples had detectable SARS-CoV-2 transcripts) (Supplementary Fig. 3). This indicates either that the vast majority of patients have cleared the virus from these organs by the time of death, or that these organs had never been infected.

**Fig. 5.**
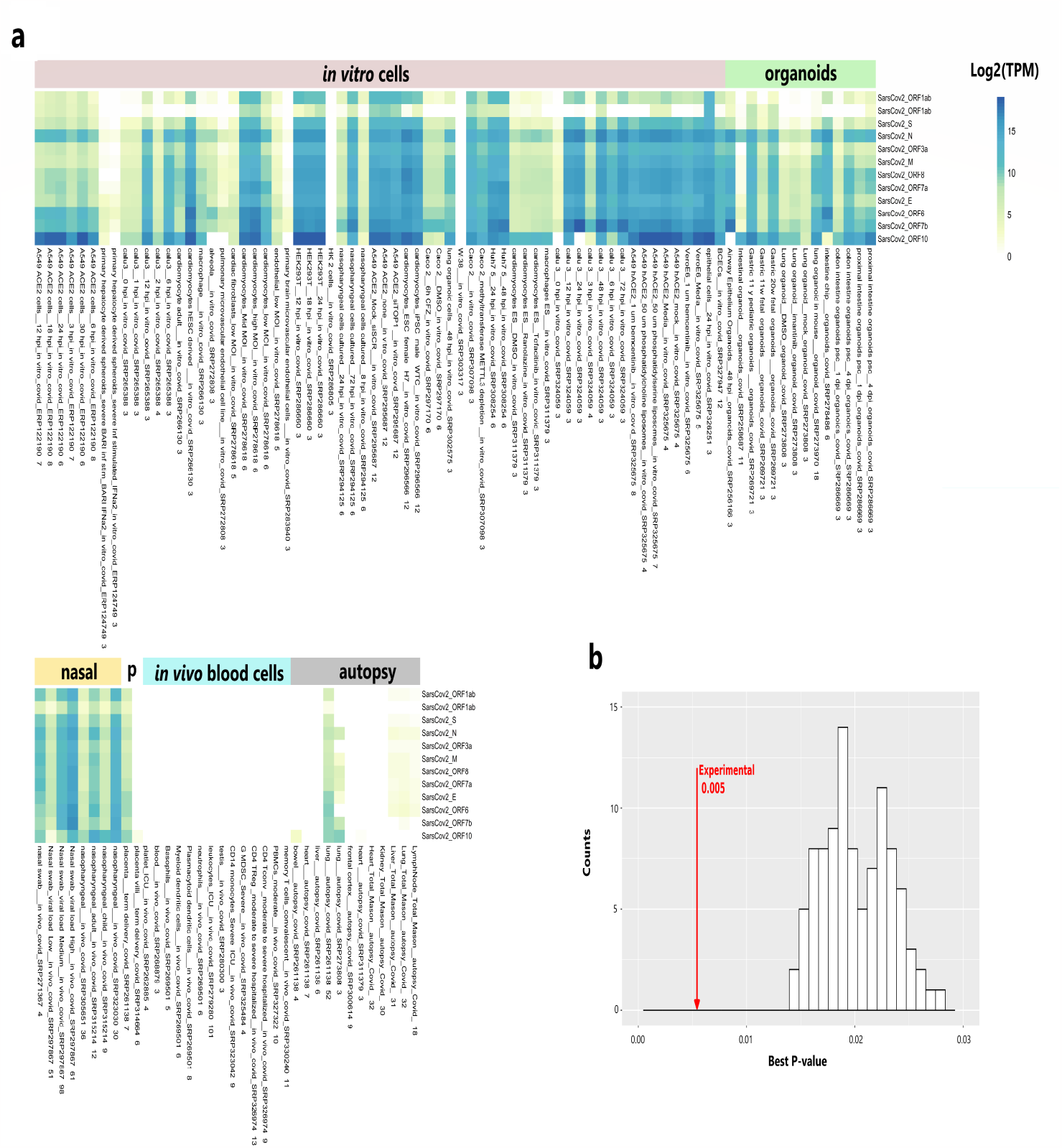
Abundance of ORF10 and other SARS-CoV-2 transcripts across multiple conditions. **A**. Heat map of diverse human tissues, cells, and organoids after SARS-CoV-2-infection. 2,070 SARS-CoV-2-associated samples from 60 independent studies were retrieved from Sequence Read Archives. To compare expression differences among genes within each sample, raw reads were processed using pyrpipe (58) and quantified as the log2 of the transcripts per million (TPM). The TPM was then averaged across specific conditions that had at least n of 3. Metadata summary for each sample cohort is in the x-axis; the number at the bottom indicates sample size. Complete TPM data and metadata, as well as TPM heatmaps of single samples are in (Supplementary File 2, Supplementary Table 4, Supplementary Fig. 3). **B**. Functional enrichment of co-expression clusters of human host genes in batch-corrected experimental data versus 100 iterations of randomized data. The method was modified from (59)).

In contrast, SARS-CoV-2 transcripts were found in lungs from deceased COVID-19 infected individuals in 4/32 samples from one study but 2/3 and 32/52 from two other study’s (Supplementary Fig. 3). That some human lungs retain viral transcripts far into the course of COVID-19 is a possible explanation of why some patients develop Post-Acute COVID-19 Syndrome (PASC).

SARS-CoV-2 transcripts accumulated following in vitro infection in most samples, and to particularly high levels (many up to Log2(TPM) 15) in SARS-CoV-2 in vitro-infected primary cardiomyocytes, nasopharyngeal cells, A549 cells (epithelial, lung carcinoma), HEK293T cells (epithelial, embryonic kidney), VeroE6 cells (epithelial, monkey kidney), Calu3 cells (epithelial, lung adenocarcinoma), and CaCo-2 cells (epithelial, colorectal adenocarcinoma) as well as in human organoid lungs transplanted to mice, human airway epithelial, and fetal and pediatric gastric, and colon and proximal intestinal organoids (Fig. 5A, Supplementary Fig. 3). SARS-CoV-2 transcripts were undetectable in in vitro-infected HK2 (proximal tubular kidney) and Wi38 diploid cells of embryonic lung) (Fig. 5A, Supplementary Fig. 3). (The absence of SARS-CoV-2 transcripts does not imply HK2 and Wi38 cells cannot be infected with the virus, but simply that they were not infected under the particular experimental conditions.)

The pattern of ORF10 accumulation differs from other SARS-CoV-2 transcripts (Fig. 5 and Supplementary Fig. 3). Specifically, ORF10 is the most abundant SARS-CoV-2 transcript in numerous samples from in vitro-infected intestinal organoid models, ACE2-A549 cells, cardiomyocytes and macrophages. In contrast, ORF10 transcript is present at very low levels or not detected in several nasopharyngeal samples from humans with COVID-19, and in multiple lung-related cells treated in vitro with SARS-CoV-2, despite other SARS-CoV-2 transcripts being more abundant (Fig. 5 and Supplementary Fig. 3).

We assessed expression of the human host genes in 3208 RNA-Seq samples, including control samples. We quantified expression of the canonical GenCode-annotated coding and non-coding genes and the highly-expressed evidence based (EB) genes (60). Mining a massive amount of RNA-Seq expression data can shed insights into host gene functions and processes integral to COVID-19 disease. To optimize biological signal in the data, and reduce noise associated with multiple batches, we gauged the effectiveness of 12 methods/parameters for data processing by creating a large pair-wise co-expression matrix for raw counts and batch corrected counts, partitioning the matrix by Markov Chain Clustering (MCL) (61) and calculating the average of the best adjusted p-values for each cluster’s Go-terms (59). For each approach, we compared the cluster enrichment of the experimental data to that of randomized data by creating clusters of the same sizes but randomly shuffling the genes throughout the clusters for 100 iterations (See Methods) (59). Clusters of the greatest biological coherence, and with the greatest number of genes in clusters, were obtained from Combat-Seq-batch-corrected data followed by a pairwise Pearson’s correlation matrix (cut-off of 0.8) (Fig. 5B) so we chose these data for further study. The MCL clusters represent functions that are altered under the experimental conditions represented in the data, covering experiments centered on SARS-CoV-2 infections across cell and tissue types. Individual MCL coexpression clusters are enriched in immune-related processes such as “positive regulation of interleukin-1 beta production”, “lymphocyte differentiation”, “monocyte differentiation”, or “defense response to virus”; mitochondrial processes such as “autophagy of mitochondrion”, “aerobic respiration”, “establishment of mitochondrial localization”, “inner mitochondrial membrane organization”, “mitochondrial gene expression”, or “mitochondrial translation”; and more (Supplemental Table 5).

### Expression of ORF10 in A549 and HEK293 cells is associated with multi-pronged changes in gene expression

Expression of ORF10 has been shown to induce mitophagy resulting in the degradation of the mitochondrial antiviral signaling protein (MAVS), antagonizing an innate immune response (62, 63). However, no one had yet looked into the effect of ORF10 expression on the expression of host transcripts. To gain insight into the overall changes in gene expression induced by ORF10, we used a non-targeted approach, generating doxycycline-inducible A549 and HEK293-T cell lines driving ORF10 expression. Because doxycycline itself can alter gene expression (64), we used doxycycline-inducible cell lines driving GFP as control. We selected for cells with no or minimal ORF10 baseline expression but robust induction following doxycycline.

RNA was isolated from A549 and HEK293-T cells (4 samples per condition) that were treated with doxycycline for 5 days. ORF10 RNA and protein accumulation were significantly increased after addition of doxycycline to the medium. Cell viability was not impacted by ORF10 expression. We quantified the levels of human transcripts in the cell samples, including all those currently annotated in GenCode and 79,203 novel Evidence-Based (EB) human transcripts, many of which have been determined to be orphan genes (60). We determined those genes that were DE associated with ORF10 expression, using a adj p-value cutoff of <= 0.05.

Genes associated with pathways of mitochondrial dysfunction and immune pathways are affected (Fig. 6A, Supplementary Table 6). ORF10 expression robustly decreases the expression of key genes involved in mito-chondrial oxidative phosphorylation (OXPHOS). The 160 OXPHOS-complex protein subunits are mostly encoded by nuclear DNA, with 13 encoded by the mitochondrial genome (mtDNA); nuclear- and mitochondrially-encoded ribosome proteins, 12S rRNAs, 16S rRNAs and 22 tRNAs are necessary for mitochondrial protein synthesis, and hence OXPHOS (66).

**Fig. 6.**
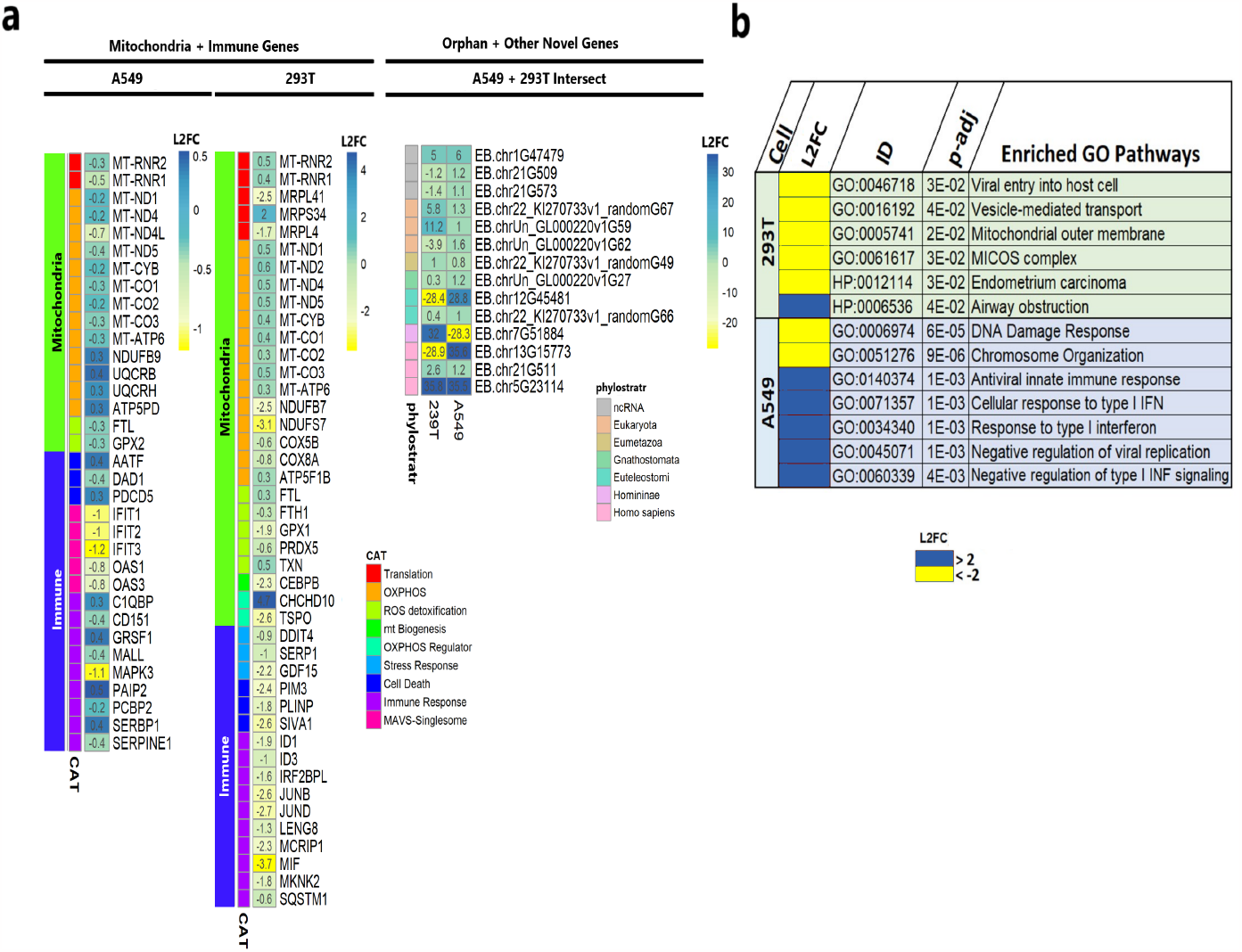
ORF10 expression induces mitochondrial and immune dysfunction. ORF10 expression was induced in A549 and 293T cells. Levels of human host GenCode-annotated and novel EB genes were quantified. Differentially expressed (DE) genes were considered as p-adj <0.05. Data for all genes is in Supplementary Table 5. **A**. Heatmap depicting Log2 fold change (L2FC) of DE mitochondrial and immune genes. CAT, functional category. Fourteen human orphans and other novel transcripts (EB) are DE across cell types. Phylostratal designation reflects the era of gene origin. ncRNA, non-coding RNA. **B**. Functional enrichment analysis of the 200 DE genes with the greatest L2FC. ToppGene (65) was used to determine GO and HP enrichment.).

Expression of ORF10 in both cell lines results in a perturbation of expression of the majority of mitochondrially-encoded genes, and induces a variety of cell-line-specific responses. In A549-cells, ORF10 decreases expression of 12S MT-RNR2 and 16S MT-RNR1 and mitochondrially-encoded genes required for the electron transport chain: complex I subunits MT-ND1, MT-ND4, MT-ND4L, MT-ND5; complex III subunit MT-CYB; complex IV subunits MT-CO1, MT-CO2, MT-CO3; and complex IV subunit MT-ATP6. In 293T-cells, expression of ORF10 results in a robust decrease in the expression of the nuclear-encoded mitochondrial ribosomal proteins MRPL4, MRPS34, MRPL41 and complex I subunits NDUB7 and NDUFS7, along with an increase in expression of additional nine mitochondrially-encoded OXPHOS genes (Fig. 6A). These alterations would disrupt mitochondrial function by decreasing OXPHOS and mitochondrial membrane potential (ΔΨm), and lead to elevated mitochondrial reactive species (mROS) production. ΔΨm reduction and mROS increase can trigger mitophagy by the host to maintain mitochondrial homeostasis (67, 68).

These data indicated that ORF10 expression induces mitochondrial dysfunction to trigger mitophagy and the degradation of MAVS to abate the immune response. Consistent with this concept, despite the absence of an immunoregulatory stimulator we observed a downward trend of innate immune genes in ORF10-expressing A549-cells and 293T cells. Down-regulated genes in A549-cells expressing ORF10 include IFIT1, IFIT3, MALL, and PCBP2; in 293T-cells expressing ORF10, MIF, JUND, PIM3, DDIT4, SIVA1 are downregulated.

To expand identification of human orphan genes that are expressed only under specific conditions, we analyzed RNA-Seq raw data from over 30,000 samples of diverse human cells, tissues and diseases, including cancers and COVID-19, and identified 79,203 previously-unannotated human transcripts having a high mean expression level under one or more biological conditions (60). Most transcripts contain ORFS predicted to encode human orphan proteins.

ORF10 expression in A549 and 293T cells results in differential expression of only tens of these 79,203 recently identified transcripts (Supplemental Table 6, adj p-value <0.05). ORF10 expression induces up-regulation of 19 transcripts and down-regulation of three in A549 cells (Supplementary Fig. 5), and the up-regulation of 13 transcripts and down-regulation of 28 transcripts in 293T cells (Supplementary Fig. 4). Interestingly, 14 of these differentially-expressed transcripts are shared between 293T and A549 cells; three of these are orphan genes-present in Homo sapiens only- and one is restricted to homininae (present in African apes and humans) (Fig. 6A). Many are up- or down-regulated by more than 10-fold. Five, such as EB.chr5G23114, are detectably expressed only under one condition or the other. Eight of the DE genes shared across both cell types are expressed in the same direction; the others are up-regulated in one cell line and down-regulated in the other (Fig. 6A). A similar difference in direction of expression of EB genes between cell or tissue types has been noted for functionally-annotated genes including genes involved in mitochondrial and/or immune processes (Fig. 6A and (69)). These transcripts provide priority targets for future research into the ORF10 mechanism of action.

## Discussion

Orphan genes constitute nearly one-third of all ORFs in many viral genomes(70); one Yaravirus genome is almost exclusively composed of orphan genes (71). In contrast, ORF10 is the only orphan gene yet identified in the genome of SARS-CoV-2; it comprises about 0.4% of the genome. ORF10 sequence has maintained high fidelity despite 7.5E+18 generations of reproduction of SARS-CoV-2 (based on each infected person carrying 10 billion virions during peak infection (72) and 650,000,000 infected humans worldwide), and on the relatively high mutation rates in RNA coronaviruses including SARS-CoV-2 (estimated as 1.3 × 10^6^ per-base per-infection cycle (73)). Thus, although SARS-CoV-2 can be transmitted without ORF10 and SARS-CoV-2 replicates without a full-length ORF10 (28), fewer than 5% of genomes have even a single mutation in ORF10 relative to the original Wuhan-Hu1reference strain, and no sequence deviations from the Wuhan-Hu-1 ORF10 sequence have persisted over time. In contrast to ORF10, ORF1a, ORF1b, ORF3a, E, M ORF6, ORF8, N and S genes have diverged significantly in NT and AA sequence; this is most evident with the recent emergence of the Omicron VOC, which has 50 new pervasive mutations, 32 in the spike protein (74, 75).

Novel proteins encoded by orphan genes provide new elements that enable innovative remodeling of evolution(5, 12– 14, 76, 77). Many proteins encoded by orphan genes of cellular organisms and virus, including ORF10, are promiscuous, binding to a range of host cell molecules (1, 18, 42– 46, 78, 79), thus potentially having a range of functions. Viral orphan genes characterized to date promote transmission, replication, and/or reproduction. Interactions among de novo orphan gene proteins of phages and those of their pseudomonad hosts create a cellular environment that is optimized for reproduction, thus enabling the virus to evade bacterial host defense systems(18). Other viral orphan gene proteins bind or influence transcriptional factors in human hosts (78), for example acting as (non-canonical) histones (79).

That only SARS-CoV-2, Pangolin-CoV-2019, and Bat-SL-CoV-RaTG13 viruses have a full-length ORF10 open reading frame is indicative of ORF10 having emerged de novo in that lineage’s common ancestor. Consistent with the *de novo* emergence of ORF10 as a protein-coding gene, the ORF10 AA sequence is highly positively selected in SARS-CoV-2, pangolin, and bat, whereas the truncated ORF10 open reading present in other SARS-CoV lineages is neutrally evolving (80).

Neither conservation of sequence nor a demonstration of ORF10 cellular action provide direct evidence as to whether the ORF10 gene is physiologically significant in human disease. We anticipated that combining genomic data and disease severity metadata from individuals with COVID-19 might reveal whether ORF10 is physiologically significant. By evaluating the clinical outcomes associated with mutations in the ORF10 gene obtained from hundreds of thousands of SARS-CoV-2-infected individuals, we show that deviations from the canonical ORF10 sequence are associated with reduced COVID-19 severity in humans. These results indicate that ORF10 function is vital for viral efficacy.

Whether ORF10 might function directly as RNA had not been experimentally addressed, however several lines of evidence support this concept. The phylogenetic conservation of sequence and structure in ORF10 RNA is consistent with a direct function for the RNA. The observed covariation in each stable hairpin of ORF 10, arising from compensatory base changes among the SARS-CoV-2 and other virulent Betacoronavirus, including bat, pangolin and SARS-CoV, is a validation of model structure. Also, the high percentage of C to T mutations among the mutant ORF10 sequences is indicative of a constraint associated with RNA (or DNA) secondary structure.

Structural aspects of the RNA are also consistent with a function for ORF10 RNA. The secondary structure of ORF10 RNA contains low (i.e., negative) ΔG z-score regions, representing sequences with a non-random nucleotide order whose nucleotides base pair to form structures with much greater stability than would be expected based on nucleotide composition. Sequences with low ΔG z-scores are likely selected for, thus maintaining a certain nucleotide sequence order for structure/function. While the average per NT ΔG z-score for the ORF10 region (−1.19) is over a full standard deviation lower than randomized sequences (of identical nucleotide composition), it is not more stable than the overall average ΔG z-score of the entire SARS-CoV-2 genome (−1.49). Indeed, low average genome z-scores appear to be a feature of the Coronaviridae family, especially when compared to other riboviruses (81). For example, the HIV and ZIKA genomes have average genome z-scores of - 0.5 (82). Given the large size of the SARS-CoV-2 genome, pressures for compact folding and packing of the RNA to fit in the viral capsid favors the genome forming a high degree of structure. In addition, several RNA structures within the SARS-CoV-2 genome have been shown to perform regulatory functions, in particular, the 5’ UTR and the frameshift stimulatory element (83).

Our finding that ORF10 transcript is highly expressed in host cells infected with SARS-CoV Betacoronavirus, which lacks the ORF10 CDS, is consistent with the transcript itself being functional. Potentially, ORF10 RNA might interact with regulatory molecules or chromatin to modulate host functions such as transcript stability, localization, or translational efficiency. The latter is perhaps more likely, considering the presence of the hairpin structures at the 3’ region of SARS-CoV-1, which does not contain an open reading frame.

Perhaps the strongest evidence that ORF10 functions as RNA is the milder COVID-10 symptoms associated with individuals infected with SARS-CoV-2 with RNA NT mutation C29659T.

The large gap we observed in the expected location of the ORF10 Stem 1 loop of seasonal Betacoronavirus and Alpha-coronavirus genomes HCoV-OC43, HCoV-HKU1, HCoV-229E, and HCoV-NL65 might relate to a loss of the function of ORF10 RNA in these human seasonal coronaviruses. Considering our findings that genetic disruptions in ORF10 RNA led to milder COVID-19 symptomatology, this absence of a full ORF10 RNA in human seasonal coronaviruses is consistent with the milder disease that may be induced by these viruses(84).

Because RNAs can form both stable, rigid structures and unstable, dynamic structures; a single RNA transcript can contain different structural landscapes. As with proteins, the stability and dynamics of a functional RNA motif are often carefully balanced for precise regulatory function. Disruptions in stability or dynamics can significantly impact the function of the motif. The clinically relevant synonymous mutations in ORF10 alter the positional entropy of the region, and this could be a factor in their effect on patient outcomes. These mutations may also affect sequence-specific binding sites for unknown trans-acting factors, and disruption of these potential binding sites could lead to a loss of interaction. The precise roles of the unusually ordered, stable and conserved ORF10 RNA structure, and how the mutation associated with reduced clinical symptoms may affect this function, is a significant target for further experimental analyses.

Successful infection and replication requires viruses to be able to attenuate innate antiviral responses (85). A characteristic feature of COVID-19 is its dysregulated immune response, with impaired type I and III IFN expression and an overwhelming inflammatory cytokine storm. RLRs, MDA5, MAVS, and cGAS–STING signaling pathways are responsible for sensing viral infection and inducing interferon (IFN) production to combat invading viruses. Consistent with this viral requirement to block host-mediated innate immune activation, ORF10 expression in A549 cells has been shown to impair cGAS–STING and MAVS signaling, thereby antagonizing innate antiviral immunity and promoting viral persistence and replication (47). Mechanistically, ORF10 protein interacted directly with STING, inhibiting STING–TBK1 association, STING oligomerization, and trafficking of STING to the Golgi (47). Our meta-analysis of RNA-Seq data shows coexpression of ORF10 transcript level with that of STING1 across multiple conditions, also consistent with a role of ORF10 in the induction of STING1 transcription or increased STING1 RNA stability.

ORF10 expression in hACE2-HELA cells (43) induced many of the gene expression changes we see in A549 cells. ORF10 abated elevation of levels of the SARS-CoV-2-induced ISG15 and OAS1 mRNAs and proteins, and decreased levels of IRF3, MAVS, TBK1, RIG-I and MDA5 proteins (43). Mechanistically, ORF10 binds and activates Nip3-like protein X (BNIPL1/NIX), inducing mitophagy, and leading to MAVS degradation, blockade of IFN responses, and promotion of viral replication(43, 47). Other viruses, including HCV (86, 87), HBV (88, 89), HPIV3 (90) and HHV-8 (90), also have been shown to trigger mitophagy, promote persistent infection and attenuate innate immune responses.

In a different cellular context, epithelial cells, ORF10 expression impaired cilia function and caused lung damage when expressed in rodent models (91). Specifically, ORF10 interacts with the ZYG11B subunit of CUL2ZYG11B protease (42, 92, 93), thereby increasing the overall E3 ligase activity, and triggering proteasome-mediated degradation of intraflagellar transport (IFT) complex B protein, IFT46 (92). Exposure of primary human nasal epithelial cells (HNECs) or the respiratory tract of hACE2-expressing mice to ORF10 results in ciliar-dysfunction, including in HNECs a rapid loss of the ciliary layer via ORF10-induced IFT46 degradation (92). These findings indicate potential functions of ORF10 in COVID-19. First, ORF10 impairs the cGAS–STING and MAVS signaling to antagonize innate antiviral immunity and promote viral persistence and replication (43, 47). Second, ORF10 interaction with the CUL2ZYG11B complex triggers the proteasome-mediated degradation of IFT46, resulting in ciliary dysfunction (92). Our pathway enrichment analysis indicates that ORF10 expression induces dysregulation of orphan genes and OXPHOS genes and is associated with airway obstruction and DNA damage response, providing new insight into how ORF10 can cause damage.

In line with ORF10 impairment of the cGAS–STING and MAVS signaling (43, 47), we hypothesize that the ORF10 sequence is conserved in part because it provides a selective advantage to viral replication by antagonizing innate antiviral immunity to promote viral persistence and replication. Consistent with this concept, silencing ORF10 via siRNA-infected Hela-hACE2 cells decreased levels of MAVS protein and viral replication (43).

ORF10 might impact COVID-19 severity by increased viral replication (43), viral persistence, and/or ciliary damage via IFT46 degradation. This could explain why mutations that affect the sequence of ORF10 protein resulted in decreased COVID-19 severity, and reflect the importance of this novel orphan gene to the fitness and deadliness of SARS-CoV-2.

Our meta-analysis quantified SARS-CoV-2 transcript accumulation across raw reads of RNA-Seq data from thousands of samples; in many of these studies SARS-CoV-2 transcripts had never been quantified. Our results are generally consistent with previous reports of SARS-CoV-2 levels determined by plaque-forming assays and RNA-Seq studies analyzed independently (69). For example, SARS-CoV-2 transcripts were not detected in autopsied kidney, heart, liver or lymph nodes of individuals who died from COVID-19, but were found at a low level in autopsied lungs of these individuals (69).

A striking finding of our meta-analysis is the disjunctive accumulation of ORF-10 RNA in relation to other SARS-CoV-2 transcripts. Notably, ORF10 transcripts are highly accumulated in intestine organoid models, whereas other SARS-CoV-2 transcripts are lower or not detected, however, a cohort of the nasopharyngeal samples have only negligible ORF10 but high levels of other SARS-CoV-2 transcripts. This phenomenon might be due to differences in ORF10 RNA stability, potentially associated with its unique stem-loop structure that might impact degradation in some cellular contexts. The biological significance to the host is a subject for future investigation. Thus, the ORF10 transcript may have a similar accumulation early in infection relative to the other transcripts, but could predominate in post-acute infected tissues due to increased stability. Consequently, increased ORF10 transcripts could aid the virus in evading an innate immune response for an extended period, resulting in increased viral persistence. This prolonged elevation of the ORF10 transcript could also contribute to long COVID. The biological significance to the host is a subject for future investigation.

The sometimes inconsistent literature on the role of ORF10 emphasizes the importance of carefully interpreting the effect of viral genes in the context of viral load, time, and tissue/cell type. For example, it was reported that the ORF10–Cullin-2–ZYG11B complex is not involved in SARS-CoV-2 infection of cultured HEK293T cells (44). However, the detailed study of (92) showed that ORF10 increased CUL2ZYG11B E3 ligase activity via a physical interaction with substrate adapter subunit, ZYG11B, and consequently altered the distribution of IFT46 in cilia, resulting in massive cilia dysfunction. The discrepancy between these results was shown to result from a difference in ORF10 expression levels (92). In another example, because ORF10 was not expressed in some cells or tissues infected with SARS-CoV-2, it was reported not to be expressed in the human host (26, 27); since, there have been reports of its expression (Our metaanalysis of SARS-CoV-2-infected tissues and cells shows ORF10 transcripts in multiple cell and tissue types, but though ORF10 is typically. In a third example, temporal changes in rates of expression of ORF10 provide a possible explanation of discrepancies in experimental findings related to ORF10 effect on SARS-CoV-2 replication. Vero 6 cells inoculated with SARS-CoV-2 mutant strains containing truncated ORF10 produced thousands-of-fold *fewer* infectious particles than a control SARS-CoV-2 strain at 24 hours post-inoculation, indicating an impact of ORF10 on viral reproduction, whereas, by 48 hours post-inoculation, the levels of infectious particles were similar (28). The expression of ORF10 or other SARS-CoV-2 transcripts during the time course of this experiment was not reported.

Taken together, our current results, experimental demonstrations of orphan genes of both viruses and cellular organisms that provide new eco-environmental opportunities (5), and the molecular characterization in cell models of ORF10’s immune-related activity(43), and STING (47), lead us to surmise that emergence of the novel protein-coding orphan gene ORF10 may have played a role in enabling the SARS-CoV-2 virus to perpetrate the pandemic that has killed seven million humans.

Orphan genes have been implicated in viral evolution (94–96), however, little concerted effort has been made to track their appearance and their potential associations with emergent diseases. Our findings emphasize the centrality of ORF10 to COVID-19. That ORF10 was mostly disregarded for years, despite the focus of the biomedical community on the pandemic, illustrates a correctable deficit in scientific approach. Routinely including both host and pathogen orphan genes in biological studies is crucial to understanding recent evolutionary trends. We advocate that mechanisms be set in place to monitor orphan genes as they arise in pathogenic viruses.

## Methods

### Expression of ORF10

ORF10 was cloned into pLVX-EF1alpha-IRES-Purovector, swapped into a TRE construct, and used to generate lentivirus. A549 and HEK293-T cells were grown for four days and then transduced and selected with puromycin to generate doxycycline-inducible cell lines. Clones were generated and allowed to grow out and samples were then tested for baseline ORF10 expression and then ORF10 induction with doxycycline. Cell lines that had no to minimal ORF10 expression at baseline and then had robust induction were used for downstream assays. RNA was isolated from A549 and HEK293-T cells that were either treated with or without doxycycline for 5 days. RNA (RIN>7.5) was used for library preparation and RNAseq was performed with NovaSeq. Cell viability was not impacted by ORF10 expression.

### Compilation and processing of COVID-19 RNA-Seq datasets for pan-tissue analysis

Bulk RNA-Seq data were downloaded from SRA via pyrpipe (58), and the corresponding sample meta-data were obtained from the SRA using https://github.com/jahaltom/GetMetaSRA. Using pyrpipe, samples were trimmed for quality and adapters and mapped using Salmon (97) to the human transcriptome (GencodeV36), SARS-COV-2(ASM985889v3) transcriptome, and human evidence based (EB) gene transcripts including novel alt-spliced, intronic, and intergenic genes (ref-Singh et. al 2023bioarchives) following the pipeline in https://github.com/jahaltom/COVID-19-Quantification. To increase mapping accuracy, the human genome along with viral decoys and spike-ins from the Genomic Data Commons (GRCh38.d1.vd1) were used as decoys for Salmon. Samples were aggregated into groups based on covid-status, tissue, treatment/other variables, study, time post infection, and in vivo/in vitro status. TPM were averaged for groups with a sample size of at least 3. The log2 of the averaged TPM was used for the heatmap.

### Gene clustering and GO analysis

Pairwise Spearman and Pearson matrices at 3 coefficient cutoffs (0.8, 0.85, and 0.9) were created from raw counts (Supplemental File 1) and from ComBatSeq batch-corrected counts using study as “batch” (Supplemental Table 4). The subsequent correlation matrices were grouped by Markov Chain Clustering (MCL) (61), filtered for clusters with ten or more annotated genes, and Go-terms (Biological Process)(98) were calculated for each cluster. For each method, Go-Terms were calculated for identical numbers of clusters and clusters of identical sizes, but with randomized gene assignments for 100 iterations (59). Randomization was done in python.

### RNA structure predictions

Data used in analysis that had been previously generated using ScanFold (82) was retrieved and downloaded from the RNAStructuromeDB database https://structurome.bb.iastate.edu/sars-cov-2. Data not already in the RNAStructuromeDB was analyzed by ScanFold and added to it. The Integrative Genomics Viewer (IGV (99)) was used to visualize data tracks, including the arc diagram (base pair) track and the per nucleotide (NT) ΔG z-score track, and used for generation of some figure elements in Fig. 4A. For the 2D structural model in Fig. 4B, - 1 z-score base pairs and lower (as modeled by ScanFold) for the ORF10 region were constrained to be paired and the entire region was refold using RNAfold to fill in less significant base pairs and/or potential longer-range interactions missed by ScanFold. The resulting model was visualized and annotated using the VARNA visualization tool (100). The per NT z-score data overlaid on the Fig. 4B model can be accessed via the FinalZavg.wig file from the downloaded SARS-CoV-2 results. Covariation analysis was previously performed on all ScanFold predicted motifs which contained at least one - 1 G z-score base pair (81). Covariation analyses were completed using the cm-builder (101) pipeline which utilizes Infernal (102) for initial sequence alignment and then R-scape (103) to determine whether covariation is statistically significant. In this study, a more detailed analysis of ORF10 was performed using data available for download from https://structurome.bb.iastate.edu/sars-cov-2. Alignment (stockholm) files for the first and second hairpins of ORF10 (Motif 523 and 524 in Dataset 3 of (81), respectively) were used to generate conservation plots (using R-chie (104)) from select viral sequences: SARS-CoV-2, SARS-CoV-1, bat, and pangolin. ScanFold and RNAFold were used to assess how synonymous mutations in ORF10 that affected clinical outcomes disrupted the secondary structure of ORF10 mRNA. Using RNAFold, we calculated the positional entropy, a measure of how likely a nucleotide is to be in a particular configuration, for the wild type sequence and each sequence with a clinically relevant synonymous mutation.

### 3D protein structure predictions

Amino acid sequences for SARS-CoV-2 mutations were obtained by computational translation from the nucleotide sequence. The AA sequences were input to RGN2 (reference) for 3D protein structure prediction. The resulting pdb files were viewed using iCn3D https://www.ncbi.nlm.nih.gov/Structure/icn3d/full.html, which allowed for an interactive exploration of each predicted protein structure, including hydrophobic/philic nature and charge distribution and solvent accessibility. Protein disorder was predicted using IUPred2A (105). Emboss (106) was used to calculate the isoelectric points. Custom python script was written to compute CDS length, GC%, Codon Usage (CU) and Relative Synonymous Codon Usage (RSCU). Atomic coordinates were aligned by the SSM superposition algorithm in Coot (55), including representation as a ribbon model and Van der Waals surface.

### Identifying ORF10 mutations from genetic sequences of SARS-CoV-2 Variants of Concern

All available sequences with associated clinical data belonging to Pango lineages B.1.1.7/Alpha variant of concern (VOC), B.1.351/Beta VOC, P.1/Gamma VOC, B.1.617.2.X/Delta VOC, and B.1.1.529.X/Omicron VOC were downloaded from GISAID database https://www.gisaid.org/ (48, 49) on October 1, 2021, except Omicron VOC sequences, which were downloaded on May 4, 2022. This included 210,101 SARS-CoV-2 ORF10 sequences for which there was associated clinical metadata (33,142 Alpha VOC, 3,533 Beta VOC, 6,599 Gamma VOC, 38,259 Delta VOC, and 128,568 Omicron VOC). The downloads were performed selecting the following GISAID EpiCoV options: i) complete, ii) low coverage excluded, and iii) with patient status. The lineage-specific sequence datasets were aligned using Nextalign CLI integrated in Nextcladec (107) with Wuhan-Hu-1 (NCBI Reference Sequence: NC_045512.2) as the reference sequence. The aligned lineage-specific datasets were trimmed to the ORF10 gene, selecting nucleotide positions 29558-29671. ORF10 lineage-specific alignments were analyzed using Nextclade (107), which allowed identification of substitution, deletions, and insertions in this gene. For comparisons of mutations across VOC, 10^3^ genomes were sampled randomly from each of the five VOCs. To account for there being only 3,533 Beta and 6,599 Gamma sequences, each VOCs sequences were tripled, then the 10^3^ genomes were sampled randomly using numpy (108).

We have also used the data made available at (51) encompassing the mutational patterns of 3.5 million SARS-CoV-2 sequences covering the Alpha, Beta, Gamma/Lambda, Delta, and Omicron VOI and VOC waves. We have investigated the rate of synonymous (S) and non-synonymous (NS) mutations per genome per site for each gene and identified where in the viral genome of the variants most of these mutations occurred with cumulative frequencies higher than 30%, 50%, and 90%.

### COVID-19 severity analysis as related to ORF10 sequence

Associations of mutations in SARS-CoV-2 ORF10 and clinical severity (Asymptomatic/Very-Mild/Mild vs Moderate/Very-Severe/Dead) were evaluated by Chi-square test using Python’s scipy.stats chi2 contingency (109). The data set was first filtered by limiting the 210,101 ORF10 sequences to the samples with clear metadata on clinical severity resulting in 181,755 ORF10 sequences for the Chi-square test.

## Supplementary Data

Supplementary data and files are available at https://drive.google.com/drive/folders/1IbcQGG20aR_znRKYxDmP147YAkQmZ9kq?usp=sharing Evidence-based novel human gene metadata is at https://github.com/urmi-21/Human_orphan_genes and is available for visualization on UCSC gene browser (Nassar LR, Barber GP, Benet-Pagès A, Casper J, Clawson H, Diekhans M, Fischer C, Gonzalez JN, Hinrichs AS, Lee BT, Lee CM, Muthuraman P, Nguy B, Pereira T, Nejad P, Perez G, Raney BJ, Schmelter D, Speir ML, Wick BD, Zweig AS, Haussler D, Kuhn RM, Haeussler M, Kent WJ. The UCSC Genome Browser database: 2023 update. Nucleic Acids Res. 2022 Nov 24;. PMID: 36420891) as a https://genome.ucsc.edu/s/jahaltom/Orphan%20Genes.

## Data Availability

All data produced in the present study are available upon reasonable request to the authors.
https://drive.google.com/drive/folders/
1IbcQGG20aR_znRKYxDmP147YAkQmZ9kq?usp=
sharing

https://drive.google.com/drive/folders/1IbcQGG20aR_znRKYxDmP147YAkQmZ9kq?usp=sharing

## ACKNOWLEDGEMENTS

We are grateful to members of the Wurtele lab and to our COV-IRT colleagues (https://www.cov-irt.org/) for helpful and stimulating discussions. We thank the patients and their families for their contributions to our study, without them, this work would not have been possible. This work is funded in part by the National Science Foundation award IOS 1546858 to ESW, “Orphan Genes: An Untapped Genetic Reservoir of Novel Traits”. R.E.S. is supported by NIH grants NIAID 2R01AI107301 and NIDDK R01DK121072 and the American Heart Association. R.E.S. is supported as Irma Hirschl Trust Research Award Scholar. W.N.M. is supported by the NIH/NIGMS through R01GM133810. D.C.W., J.G. and J.H. are supported by DOD W81XWH-21-1-0128 and Bill Melinda Gates Foundation Grant INV-046722 awarded to D.C.W. This work used the Extreme Science and Engineering Discovery Environment (XSEDE) supported by National Science Foundation ACI-1548562. In particular, it used the Bridges HPC environment through TG-MCB190098 and TG-MCB200123 to ESW, US and JH. The opinions expressed in this article are those of the authors and do not reflect the view of the National Institutes of Health, National Science Foundation, the Department of HHS, or the U.S. government.

